# Medium-term scenarios of COVID-19 as a function of immune uncertainties and chronic disease

**DOI:** 10.1101/2023.03.08.23287004

**Authors:** Chadi M. Saad-Roy, Sinead E. Morris, Rachel E. Baker, Jeremy Farrar, Andrea L. Graham, Simon A. Levin, Caroline E. Wagner, C. Jessica. E. Metcalf, Bryan T. Grenfell

**Affiliations:** Lewis-Sigler Institute for Integrative Genomics, Princeton University; Miller Institute for Basic Research in Science, University of California, Berkeley; Department of Integrative Biology, University of California, Berkeley; Department of Pathology and Cell Biology, Columbia University Medical Center; Department of Epidemiology, Brown University; Institute at Brown for Environment and Society, Brown University; The Wellcome Trust; Department of Ecology and Evolutionary Biology, Princeton University; Department of Bioengineering, McGill University; School of Public and International Affairs, Princeton University

## Abstract

As the SARS-CoV-2 trajectory continues, the longer-term immuno-epidemiology of COVID-19, the dynamics of Long COVID, and the impact of escape variants are important outstanding questions. We examine these remaining uncertainties with a simple modelling framework that accounts for multiple (antigenic) exposures via infection or vaccination. If immunity (to infection or Long COVID) accumulates rapidly with the valency of exposure, we find that infection levels and the burden of Long COVID are markedly reduced in the medium term. More pessimistic assumptions on host adaptive immune responses illustrate that the longer term burden of COVID-19 may be elevated for years to come. However, we also find that these outcomes could be mitigated by the eventual introduction of a vaccine eliciting robust (*i.e*. durable, transmission-blocking and/or ‘evolution-proof’) immunity. Overall, our work stresses the wide range of future scenarios that still remain, the importance of collecting real world epidemiological data to identify likely outcomes, and the crucial need for the development of a highly effective transmission-blocking, durable, and broadly-protective vaccine.

## Introduction

The emergence of the severe acute respiratory syndrome coronavirus 2 (SARS-CoV-2) has led to the ongoing, multi-year COVID-19 pandemic. Due to sustained transmission and the emergence of novel variants, this outbreak remains a public health emergency and continues to exert a significant burden across the world [1]. A number of safe and effective vaccines have been developed and deployed (e.g. [2, 3, 4]), and progress toward more broadly protective immunization continues (*i.e*. pan-coronavirus/sarbecovirus vaccines) [5,6,7]. With development and wide dissemination of effective, transmission-blocking vaccination, community immunity could prevent local transmission [8, 9, 10]. However, the rapid spread of the Omicron (and BA.2) variant among vaccinated individuals illustrates that community immunity is unlikely to be achieved via existing vaccines. Furthermore, unequal vaccine distribution and incomplete uptake are still a pressing issue, with potential consequences that include the emergence of novel variants in addition to elevated disease burden [11].

During the SARS-CoV-2 pandemic, it has become clear that we must better gauge the impact of accumulating immunity on susceptibility, the evolution of new variants, and the clinical features of subsequent infections. With simple mathematical models of immuno-epidemiological dynamics, we showed that the medium- and long-term population-level landscapes of immunity and infection crucially depend on the strength and duration of immunity following infection or vaccination [8]. We also explored the intricacies that emerge from vaccine dosing regimes (for two-dose vaccines) [10] or from the invasion dynamics in largely vaccinated populations [12], in addition to the consequences of vaccine nationalism [11]. Throughout, we have shown how epidemiological and/or evolutionary outcomes hinge on the characteristics of host immune responses.

While the mass of data generated during the pandemic (*e.g*. see [1] for a comprehensive review) has clarified many outstanding epidemiological questions, there are still major uncertainties (*e.g*. see [13]). In particular, it is crucial to determine how clinical and transmission-blocking immunity changes with repeated infection or vaccination, the impact of new immune escape variants, and the potential dynamics of long COVID. For example, widespread vaccination in many countries has blunted the impact of COVID-19 on hospital capacities (e.g. [14,15]). However, infections that are less severe (and even classified initially as “mild”) can lead to long-term debilitation [16, 17], such as neurological impact [18]. As many jurisdictions world-wide relax restrictions to return to pre-pandemic levels of interaction, the specter of elevated population-wide chronic disease following infection (‘long COVID’) looms large. Furthermore, the potential for rapid spread of novel variants has been exemplified by previous and ongoing rises in infection levels (in certain countries) due to new variants (*e.g*. previous increases driven the BA.2 variant [19]). In light of these underlying uncertainties and to inform future policy decisions, it is important to examine ranges of potential medium-term scenarios. In this work, we use simple immuno-epidemiological models to explore these issues and qualitatively examine the population-level outcomes under various scenarios, spanning optimistic to pessimistic assumptions.

We begin by extending previous modelling work [8] (see also [20]) to include characteristics of tertiary and quaternary infections. Our model extension separates individuals based upon “exposures” to SARS-CoV-2, whether by vaccination or infection (Fig. 1A). The immuno-epidemiological dynamics are schematically depicted in Figure 1B. We qualitatively capture the potential decreases in relative susceptibility to infection as host immunity slowly accumulates due to previous exposures (Fig. 1C) (hereafter referred to as the “accumulation of immunity”).

**Figure 1:**
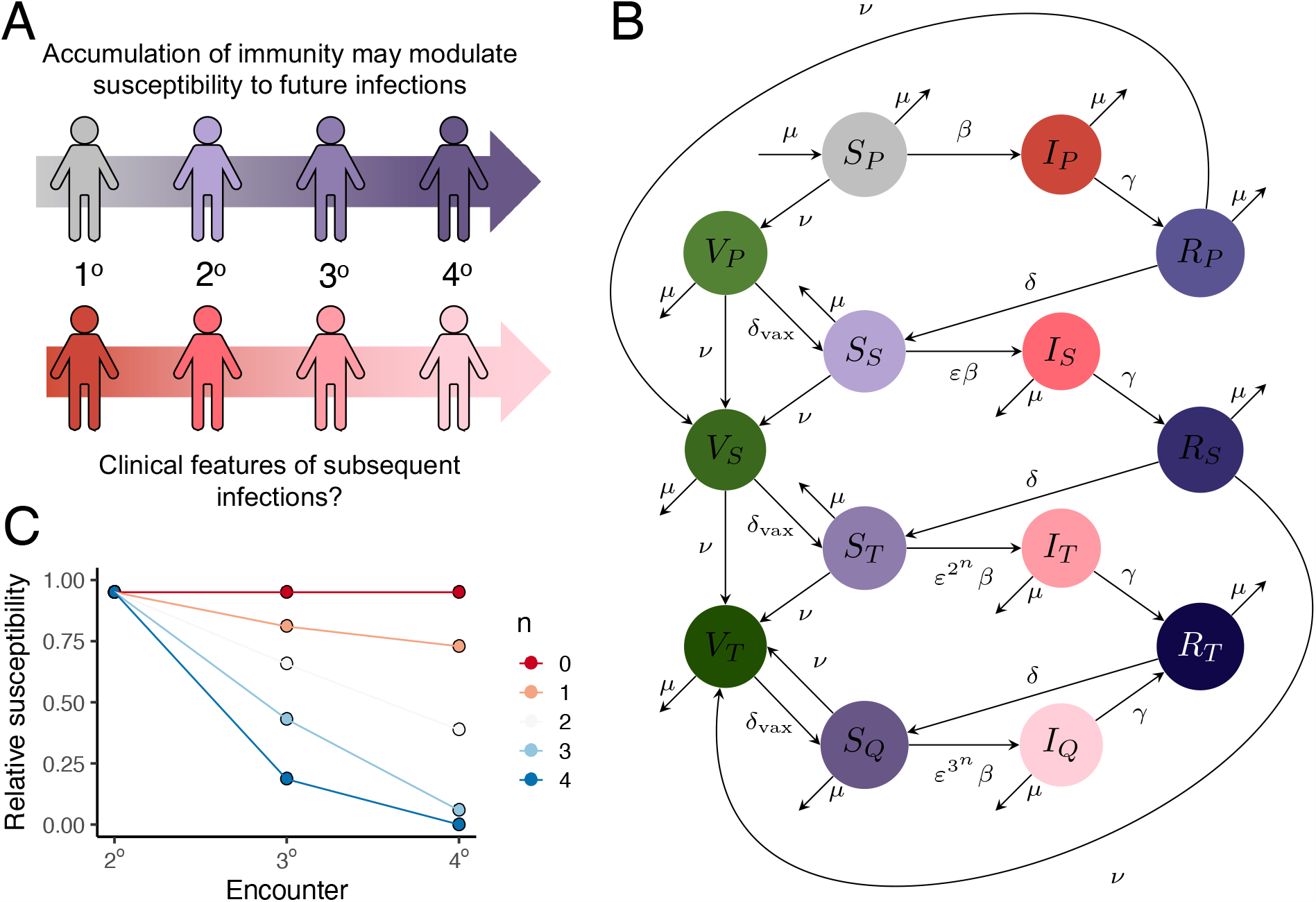
Immuno-epidemiological model schematics. (*A*) Illustration of accumulating immunity and its potential impact on subsequent infections. (*B*) Model flow diagram, modified from [8] to include tertiary and quaternary infections, and multiple vaccinated classes. (*C*) Illustrations of a range of assumptions about immunity with each infection via a change in relative susceptibility. In our model, the relative susceptibility to a secondary, tertiary, and quaternary (and beyond) infection is *ε*, 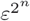, and 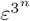, respectively. In panel *C*, we depict values of *n* ≥ 0, with larger values of *n* indicating more rapid accumulation of immunity (*i.e*. relative susceptibility to tertiary and quaternary infection are increasingly smaller).

## Results and Discussion

With this refined model, we first explore impacts of the accumulation of clinical and transmission-blocking immunity with (antigenic) exposure (infection or vaccination) on transmission, immune, and Long COVID dynamics. Second, we examine the impact of immune escape variants on population-level dynamics. Finally, we investigate the impact of decreasing immunity against tertiary and quaternary (and further) infections (*i.e*. an increase in relative susceptibility to reinfection compared to that for a secondary infection).

### Accumulation of immunity with subsequent infections or vaccinations

To examine the interactions between accumulation of immunity and reduction in susceptibility to secondary infection, we begin by exploring synoptic medium-term immuno-epidemiological landscapes. Guided by emerging evidence, we assume that the duration of complete natural and vaccinal immunity is short (see *Supplementary Materials*), and examine a range of possibilities for the accumulation of immunity and the susceptibility to secondary infection after complete immunity has waned. We also assume two periods of nonpharmaceutical interventions that reduce transmission early on, separated by a short relaxation period (*i.e*. reduction of seasonal transmission rates to 60% of its value, during weeks 16 to 55 and weeks 62 to 79).

Pessimistically, if no immunity accumulates (*i.e*., exposures beyond the first do not further decrease susceptibility to infection), small changes in the relative susceptibility to secondary infection can have large effects on the size of the second peak and can affect the depths of seasonal troughs. In line with intuition, such small changes do not have appreciable longer-term impacts on peak infection levels or their timing (Fig. 2, *top row*).

**Figure 2:**
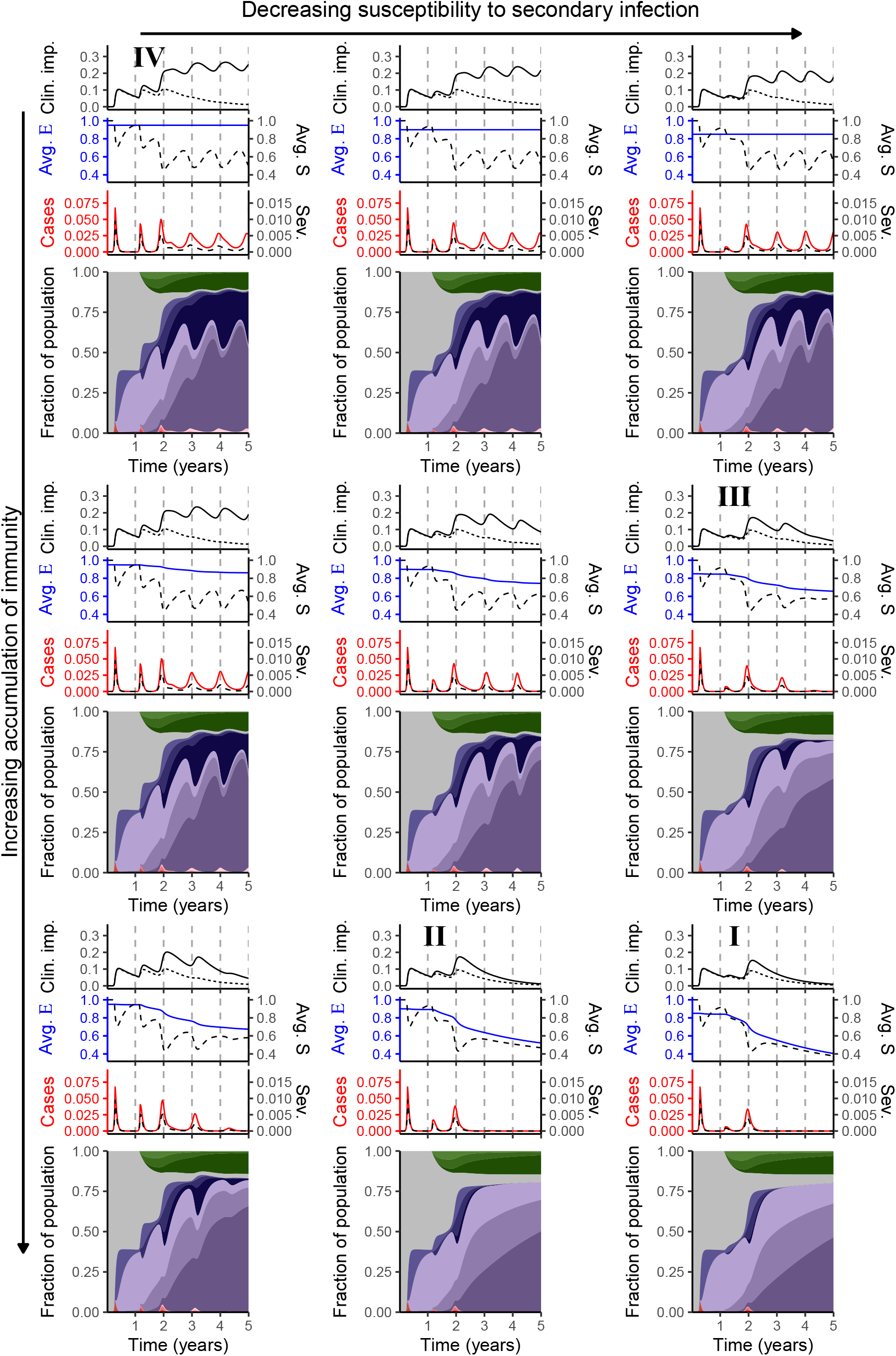
Synoptic medium-term immuno-epidemiological landscapes as functions of accumulation of immunity and relative susceptibility to secondary infection. For the panels in the *top, middle*, and *bottom rows, n* = 0, *n* = 1 and *n* = 2, respectively. In the *left, middle*, and *right columns, ε* = 0.95, *ε* = 0.9, and *ε* = 0.85, respectively. Each individual panel consists of an area plot (*bottom*) that depicts the immunological and infection status of the population. The color coding of this panel is as in Figure 1B. Above this panel are three plots: the first depicts cases (*solid red line*) and severe cases (*dashed black line*) (see *Supplementary Materials*), the second shows the average relative susceptibility to subsequent infection (*solid blue line*) and the average susceptibility (*dashed black line*) (see *Supplementary Materials* for the equations), and the third depicts scenarios for Long COVID, assuming that 30% of primary infections lead to Long COVID for on average a year (see *Supplementary Materials* for details). The *dashed line* represents an optimistic scenario where the likelihood decreases substantially (by 90%) after each exposure (up to quaternary), pessimistic, whereas the *solid line* depicts a pessimistic scenario where the likelihood decreases negligibly (by 10%) after each exposure (up to quaternary).

As the accumulation of immunity increases, the timing and magnitude of future peaks is increasingly affected by small reductions in susceptibility to secondary infection (Fig. 2, compare *bottom row* with *top row*). In turn, these changes lead to substantial differences in the number of clinically important long term infections (Long COVID, ‘Clin. Imp.’) in the *top plot* of each panel. (Note that this fraction is distinct from the more acute clinically severe fractions of infections (‘Sev’, *dashed line*) depicted above the area plot of each panel). To understand these dynamics, it is useful to examine the average relative susceptibility to subsequent infection in the population, denoted Avg. E. For weeks where there are individuals with partial susceptibility (*i.e. S*_*S*_(*t*) + *S*_*T*_ (*t*) + *S*_*Q*_(*t*) *>* 0), Avg. E is the weighted average of subsequent relative susceptibility to infection, with weights corresponding to the fraction of individuals in each partially susceptible class (see *Supplementary Materials* for additional details).

Intuitively, the drastic effects observed in Figure 2 emerge because Avg. E decreases sharply if immunity accumulates rapidly. In the *Supplementary Materials*, we further explore the combination of accumulating immunity, total cases, and clinical severity (see Figure S1 and associated discussion in *Supplementary Materials*). Additionally, the accompanying online interactive application at https://grenfelllab.shinyapps.io/chronicCOVID/ allows for further explorations.

To summarize Figure 2, we have selected four representative scenarios for the range of potential outcomes (identified as **I, II, III**, and **IV** on Fig. 2).

- Scenario **I** represents the most optimistic case: rapid accumulation of immunity, combined with a strong reduction in susceptibility following the first exposure. This combination leads to strong population immunity (*i.e*., low average susceptibility) and thus very few cases once the third peak has abated. While this Scenario is very unlikely to be attained at this stage unless more robust vaccines (or possibly antivirals) emerge, we include this scenario for comparison purposes.
- Scenarios **II** and **III** depict more pessimistic situations where either immunity accumulates rapidly but a primary exposure only slightly decreases susceptibility to secondary infection (**II**) or immunity accumulates more slowly but a primary exposure substantially decreases susceptibility (**III**). Under either of these conditions, a sizable fraction of the population experience a clinically important ‘Long COVID’ infection (*top plot*), but the magnitudes of the later peaks of infections are attenuated.
- Scenario **IV** represents a relatively pessimistic outcome with yearly seasonal outbreaks and a large fraction of the population experiencing a clinically important ‘Long COVID’ infection in the medium term (*top plot*).

### Immuno-epidemiological impacts of immune escape variants

As illustrated by the recent rapid spread of Omicron, novel variants with various immune-escape characteristics may emerge. To qualitatively examine the impact of a variant on population-level immuno-epidemiology, we simulate the emergence of a variant with no decrease in susceptibility to secondary infection, which also corresponds to no accumulation of transmission-blocking immunity with subsequent infections. We assume that the variant arises after the third year following the onset of the COVID-19 pandemic.

In Figure 3 (*top and middle rows*), we examine the potential synoptic landscapes with the spread of an immune-escape variant after week 156. These landscapes illustrate that both the rapidity with which immunity accumulates and the relative susceptibility to secondary infection before the immune-escape variant spreads can have important impacts on the size and timing of post-emergence peaks. Additionally, the burden of long COVID may be substantial.

**Figure 3:**
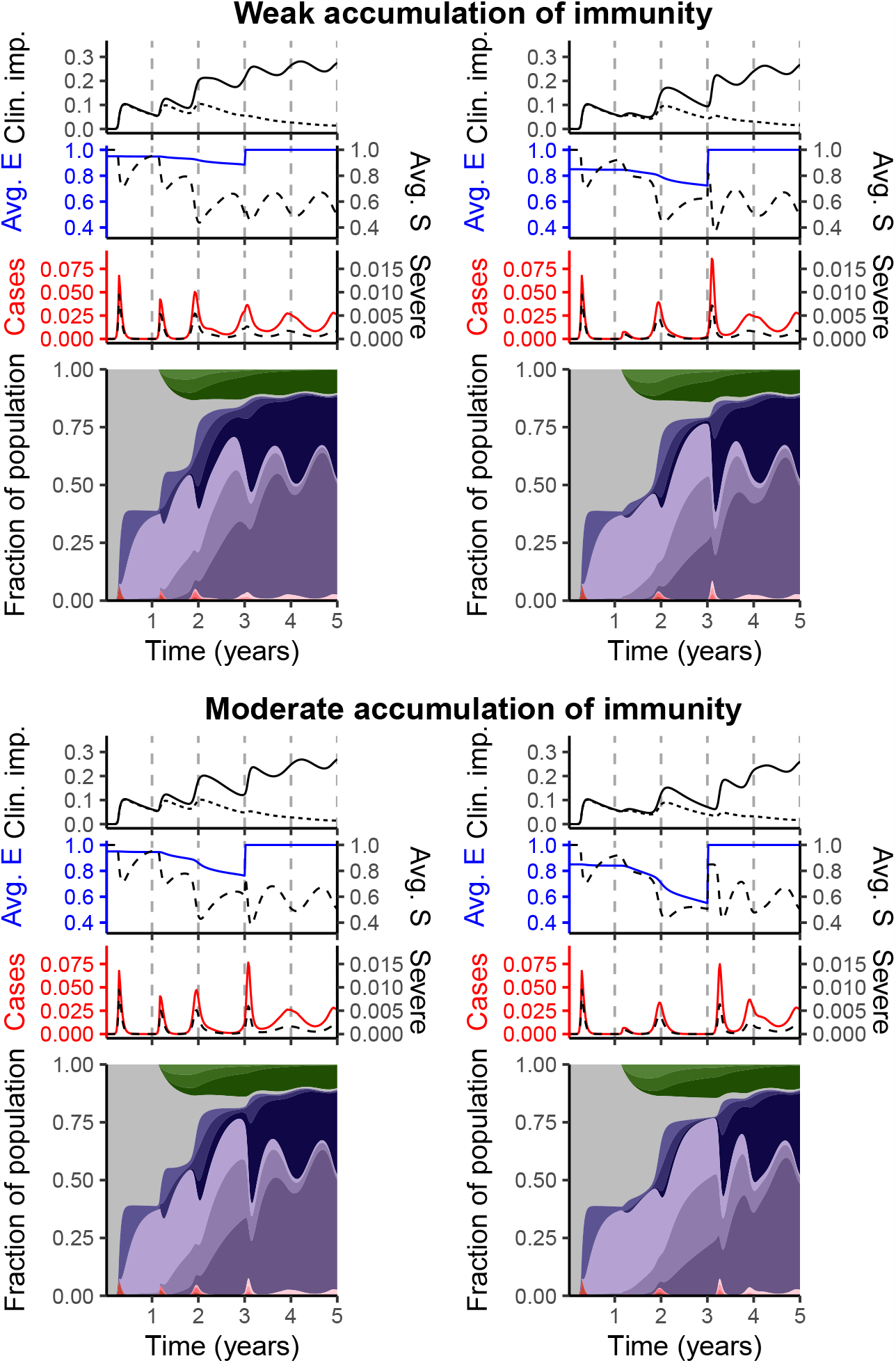
The impact of an immune-escape variant on medium-term immuno-epidemiological dynamics. To model an immune-escape variant, we assume that *ε* = 1 after week 156. The panels are as in the panels of Figure 2, selected to show the difference between weak (*i.e. n* = 1) and moderate (*i.e. n* = 2) accumulation of immunity prior to the emergence of the immune-escape variant (*top* and *bottom rows*, respectively), and for *ε* = 0.95 (*left column*) and *ε* = 0.85 (*right column*).

The impact of an immune-escape variant is especially notable when it emerges in a setting where the current variant induces a strong reduction in relative susceptibility after infection, *i.e*. reinfection is more difficult (compare *left panels* with *right panels* in Figure 3). Intuitively, this effect is due to an important increase in population susceptibility - and a corresponding rise in average susceptibility to subsequent infection (Avg. E) - when the immune-escape variant emerges.

### Decreasing immunity with accumulation of infections

A number of complexities regarding cross-protective natural and vaccinal immunity to subsequent infections can arise (*e.g*. see [21]). In a very pessimistic case, while a first infection may provide partial protection against a second infection, the combination of new variants and complexities surrounding immune responses could then increase the susceptibility to tertiary and quaternary infections. Our framework can easily incorporate this possibility through *n <* 0 (*cf*. Figure 1 and see *Supplementary Materials*).

In Figure 4, we illustrate the range of possibilities for this pessimistic setting. For simplicity, we assume that while immunity against infection may be poorer, subsequent infections still increase protection against ‘Long COVID’. While the average relative susceptibility to infection (Avg E, *blue line*) reaches 1 more rapidly as immunity is poorer, the qualitative dynamics are very similar across this range (compare rows of Fig. 4). More apparent differences begin emerging once the susceptibility to secondary infection is low (*leftmost column*, Fig. 4): poorer immunity with accumulating infections can lead to elevated infections and decreases the depths of the seasonal troughs (compare *top rightmost panel* with *bottom rightmost panel* of Fig. 4). Thus, these potential immunological complexities have a more moderate impact on medium-term outcomes even as clinical outcomes might deteriorate.

**Figure 4:**
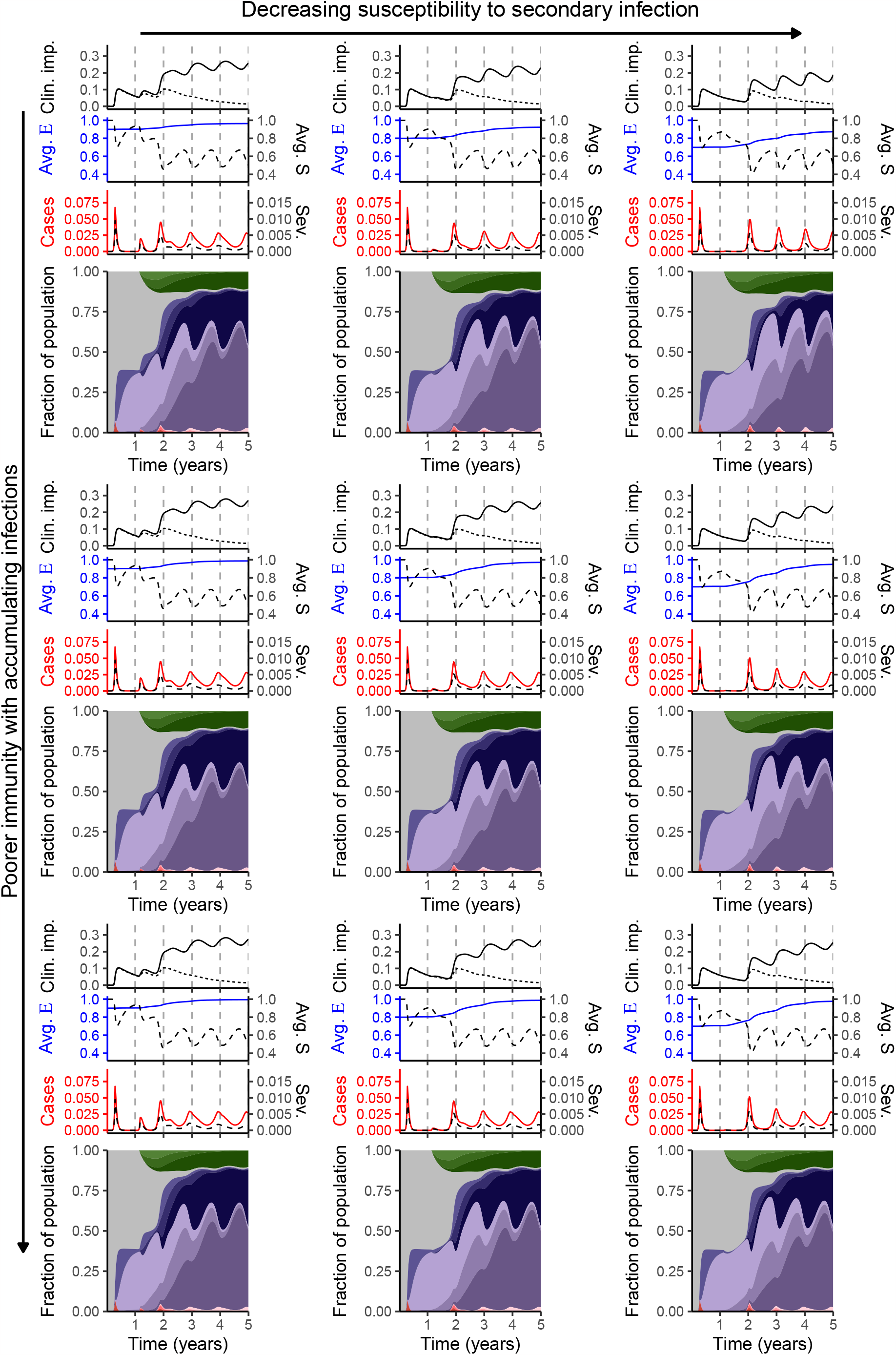
Synoptic medium-term immuno-epidemiological landscapes for a very pessimistic case where immunity is progressively worse as an individual accumulates infections, the *top, middle*, and *bottom rows* corresponding to *n* = −1,*n* = −2, and *n* = −3, respectively. The *left, middle*, and *right* columns correspond to *ε* = 0.9, *ε* = 0.8, and *ε* = 0.7, respectively. The plots within each panel are as in Figure 2.

### The development of a more robust vaccine

There are currently multiple vaccines in development (*e.g*. [5, 6, 7]) that aim to protect against a greater range of coronaviruses, including potential future SARS-CoV-2 variants. If successful, such a vaccine could generate robust immunity against the evolving SARS-CoV-2 for a much longer duration than that elicited by current vaccines. Our modelling framework allows us to explore the dynamical implications that would arise from the deployment a highly effective vaccine.

In Figure 5, we examine the impact of a very robust vaccine on immuno-epidemiological dynamics and potential Long COVID trajectories for pessimistic scenarios with poor natural and current vaccinal immunity and weak accumulation of immunity following repeated exposures. Intuitively, a robust vaccine (giving complete immunity for on average 2 years following any dose) leads to substantial decreases in infection levels and Long COVID burden (compare Fig. 5 with *middle row*, Fig. 2). If the reduction in relative susceptibility to secondary infection is strong enough, then a robust vaccine would prevent subsequent peaks in the medium term (*leftmost panel*, Fig. 5). In contrast, if the reduction in relative susceptibility to secondary infection is more modest, a small infection peak can be expected in the medium term (*rightmost panel*, Fig. 5). Echoing [8], these observations highlight the importance of rapid robust vaccine development in conjunction with using real world epidemiological data to quantify the relative susceptibilities to subsequent infections.

**Figure 5:**
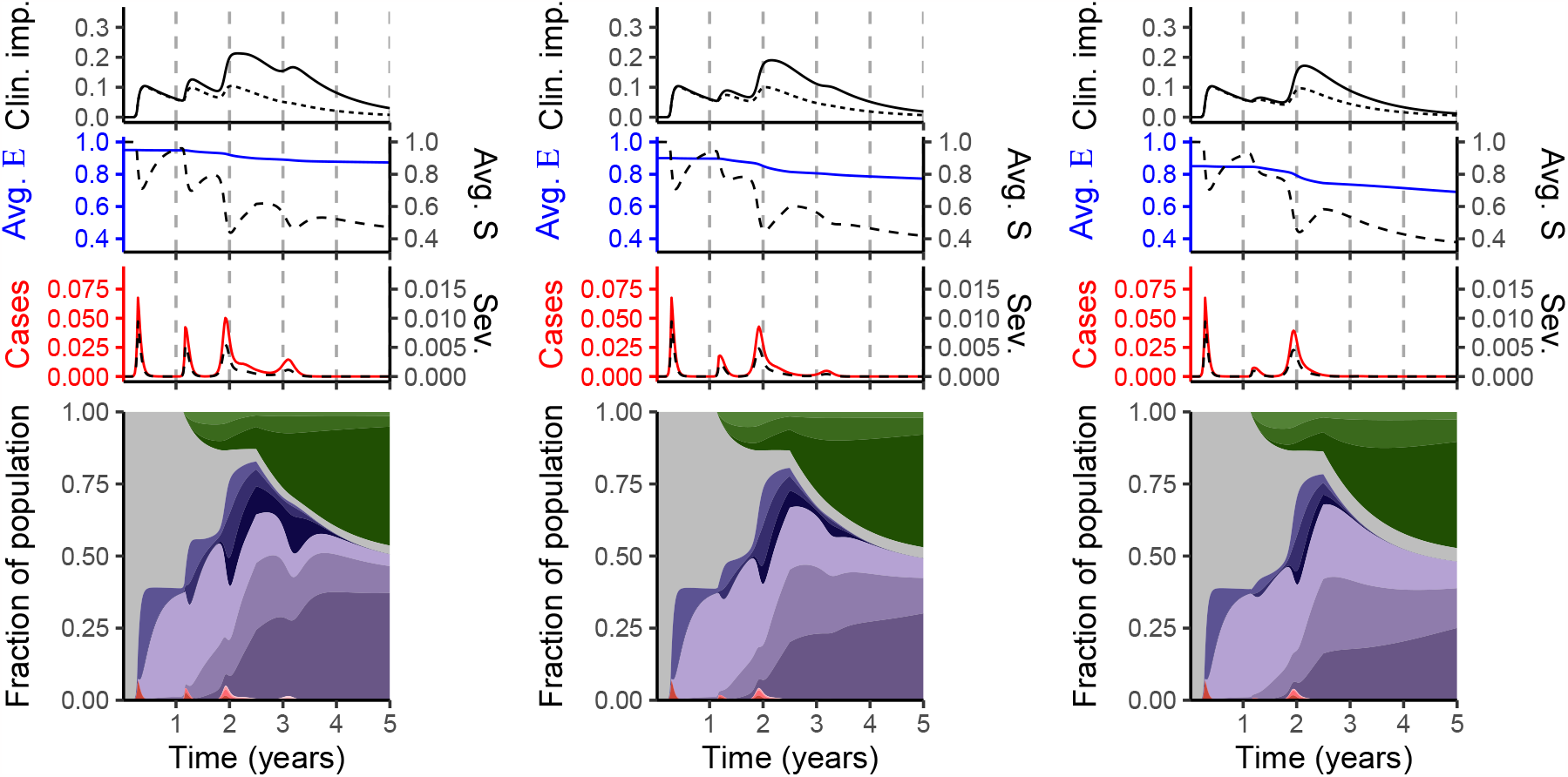
Dynamical impacts following the introduction of a robust vaccine after week 130. Here, we use scenarios with weak accumulation of immunity (*middle row*, Figure 2) and assume that vaccinal immunity conferred following any dose is 2 years (in contrast to 33% of a year for the previous vaccine). As with Figure 4, the separate stacked plots in each panel are described in the caption to Figure 2.

## Caveats and future directions

We have made a number of simplifying assumptions. First, we have focused on immune heterogeneities and omitted others, such as those arising from age [22, 23] or transmission [24, 25]. However, in previous work [8], we have shown that additional heterogeneities do not qualitatively affect the dynamics in our models. Nevertheless, for quantitative predictions of transmission and disease dynamics, future work should focus on incorporating various sources of heterogeneities in our simple immuno-epidemiological models.

Second, we have assumed that quinary encounters (*i.e*. fifth exposures) and beyond are equivalent to quaternary encounters, and that they lead to no additional accumulation of immunity. As the pandemic progresses and more data becomes available, the importance of subsequent infections will become clear, and the model could be refined accordingly. Additionally, we have assumed a simple functional form to account for the accumulation of immunity. When appropriate data become available, it will be imperative to determine, via careful quantitative calibrations, the degree to which susceptibility changes following each encounter.

Third, we have assumed throughout that relative susceptibility to subsequent infections is constant or decreases for the second infection (*ε* ≤ 1). In reality, complex interactions between prior immunity and emerging variants could potentially lead to increases in susceptibility immediately (*e.g*. antibody interference for influenza [26]). For a disease where susceptibility increases even after the first exposure, our model framework would apply by setting *ε >* 1.

Fourth, we have assumed seasonal climate-driven transmission rates from HKU1 for NYC [27]. Since emerging variants have increased transmissibility relative to the original virus (*e.g*. [28]), our illustrative results are conservative. Furthermore, as in [8, 10, 11], we have assumed simple scenarios for nonpharmaceutical interventions (NPIs).

The online application (https://grenfelllab.shinyapps.io/chronicCOVID/) can be used to examine the impacts of transmission in different climates, varying intensity and periods of NPIs, and different assumptions about host immune responses.

Finally, we have taken a very simple approach to model the emergence of an immune escape variant. As more variants emerge that differ in their capacity to evade existing immune responses, a number of complexities are likely to arise from these interactions. Building on our work, further models that carefully examine multi-strain dynamics with immune uncertainties will be crucial. In particular, it is important to understand the fate of variants, and especially current co-circulation of Omicron sub-variants. Additionally, the development and deployment of broader vaccines [5, 6, 7] will undoubtedly shape immune landscapes and the dynamics of emerging variants. Future work is needed to examine the population-level dynamical impact of such vaccines, and our immuno-epidemiological framework is a starting point for these important investigations.

## Conclusion

As jurisdictions worldwide have relaxed public health measures, and new SARS-CoV-2 variants have emerged, increases in transmission have been widely observed. However, especially in places with widespread vaccination, acute severe disease appears to have been largely blunted by prior immunity. At the same time, the appearance of multiple variants of concern since the initial emergence of SARS-CoV-2 raises the possibility of variants capable of escaping these existing immune responses from vaccination or prior infection. Additionally, the resulting dynamics of Long COVID remain worrying, and these will undoubtedly shape the longer term burden of COVID-19.

While remaining COVID-19 unknowns have considerably narrowed since early modelling work [8], we have illustrated through our model extensions that the range of potential scenarios remains substantial. As the pandemic progresses, the impacts of subsequent infection and vaccination on immune life history will become clear. In parallel, the underlying drivers of SARS-CoV-2 evolutionary trajectories may be determined (*e.g*. see [29]). As we have shown here, the combination of these factors will influence the medium-term synoptic landscape of immunity and active infections, and the burden of Long COVID. Our work highlights the importance of immunological monitoring at the population-level (see proposals for “Global Immunological Observatory” [30, 31, 32]). Used in conjunction with our models, these data would untangle the current uncertainties in medium- and long-term outcomes and thus allow for proper post-pandemic preparedness. Pessimistic scenarios of future acute and chronic disease burden also underline the high priority of maintaining momentum in developing new vaccines (notably mucosal vaccines to reduce infection; [33, 34]) and cocktails of antiviral therapies. However, production of broadly protective vaccines is not enough: achieving global dissemination, and persuading populations to accept vaccination, are crucial priorities for global health.

## Data Availability

The manuscript does not include new data.

## Acknowledgments

This work was funded in part by the Natural Sciences and Engineering Research Council of Canada through a Postgraduate Scholarship-Doctoral (C.M.S.-R.); a Charlotte Elizabeth Procter Fellowship of Princeton University (C.M.S.-R); a Miller Research Fellowship from the Miller Institute for Basic Research in Science (C.M.S.-R.); the James S. McDonnell Foundation 21st Century Science Initiative Collaborative Award in Understanding Dynamic and Multi-scale Systems (S.A.L.); the C3.ai Digital Transformation Institute and Microsoft Corporation (S.A.L.); a gift from Google, LLC (S.A.L.); the National Science Foundation (CNS-2027908, CCF1917819) (S.A.L.); and Flu Lab (B.T.G.).

## Notes

### Competing Interest Statement

The authors have declared no competing interest.

## References

[1] K Koelle, MA Martin, R Antia, B Lopman, NE Dean, The changing epidemiology of SARS-CoV-2. Science 375, 1116–1121 (2022).

[2] LR Baden, et al., Efficacy and safety of the mRNA-1273 SARS-CoV-2 vaccine. New England Journal of Medicine 384, 403–416 (2021) PMID: 33378609.

[3] FP Polack, et al., Safety and efficacy of the BNT162b2 mRNA Covid-19 vaccine. New England Journal of Medicine 383, 2603–2615 (2020) PMID: 33301246.

[4] M Voysey, et al., Safety and efficacy of the ChAdOx1 nCoV-19 vaccine (AZD1222) against SARS-CoV-2: an interim analysis of four randomised controlled trials in Brazil, South Africa, and the UK. The Lancet 397, 99–111 (2021).

[5] HAD King, et al., Efficacy and breadth of adjuvanted SARS-CoV-2 receptor-binding domain nanoparticle vaccine in macaques. Proceedings of the National Academy of Sciences 118, e2106433118 (2021).

[6] DR Martinez, et al., Chimeric spike mRNA vaccines protect against Sarbecovirus challenge in mice. Science 373, 991–998 (2021).

[7] AA Cohen, et al., Mosaic RBD nanoparticles protect against challenge by diverse sarbecoviruses in animal models. Science 377, eabq0839 (2022).

[8] CM Saad-Roy, et al., Immune life history, vaccination, and the dynamics of SARS-CoV-2 over the next 5 years. Science 370, 811–818 (2020).

[9] CM Saad-Roy, SA Levin, CJE Metcalf, BT Grenfell, Trajectory of individual immunity and vaccination required for SARS-CoV-2 community immunity: a conceptual investigation. Journal of The Royal Society Interface 18, 20200683 (2021).

[10] CM Saad-Roy, et al., Epidemiological and evolutionary considerations of SARS-CoV-2 vaccine dosing regimes. Science 372, 363–370 (2021).

[11] CE Wagner, et al., Vaccine nationalism and the dynamics and control of SARS-CoV-2. Science 373, eabj7364 (2021).

[12] CM Saad-Roy, et al., Vaccine breakthrough and the invasion dynamics of SARS-CoV-2 variants. medRxiv, 10.1101/2021.12.13.21267725 (2021).

[13] CM Saad-Roy, CJE Metcalf, BT Grenfell, Immuno-epidemiology and the predictability of viral evolution. Science 376, 1161–1162 (2022).

[14] MW Tenforde, et al., Association Between mRNA Vaccination and COVID-19 Hospitalization and Disease Severity. JAMA 326, 2043–2054 (2021).

[15] LJ Abu-Raddad, et al., Effect of mRNA Vaccine Boosters against SARS-CoV-2 Omicron Infection in Qatar. New England Journal of Medicine, DOI: 10.1056/NEJMoa2200797 (2022).

[16] JL Hirschtick, et al., Population-Based Estimates of Post-acute Sequelae of Severe Acute Respiratory Syndrome Coronavirus 2 (SARS-CoV-2) Infection (PASC) Prevalence and Characteristics. Clinical Infectious Diseases 73, 2055–2064 (2021).

[17] M Michelen, et al., Characterising long COVID: a living systematic review. BMJ Global Health 6, e005427 (2021).

[18] E Xu, Y Xie, Z Al-Aly, Long-term neurologic outcomes of COVID-19. Nature Medicine (2022).

[19] J Fonager, et al., Molecular epidemiology of the SARS-CoV-2 variant Omicron BA.2 sub-lineage in Denmark, 29 November 2021 to 2 January 2022. Eurosurveillance 27, 2200181 (2022).

[20] SE Morris, et al., Demographic buffering: titrating the effects of birth rate and imperfect immunity on epidemic dynamics. Journal of The Royal Society Interface 12, 20141245 (2015).

[21] CJ Reynolds, et al., Immune boosting by B.1.1.529 Omicron depends on previous SARS-CoV-2 exposure. Science 377, eabq1841 (2022).

[22] JS Lavine, ON Bjornstad, R Antia, Immunological characteristics govern the transition of COVID-19 to endemicity. Science 371, 741–745 (2021).

[23] NG Davies, et al., Age-dependent effects in the transmission and control of COVID-19 epidemics. Nature Medicine 26, 1205–1211 (2020).

[24] K Sun, et al., Transmission heterogeneities, kinetics, and controllability of SARS-CoV-2. Science 371, eabe2424 (2021).

[25] R Laxminarayan, et al., Epidemiology and transmission dynamics of COVID-19 in two Indian states. Science 370, 691–697 (2020).

[26] W Ndifon, NS Wingreen, SA Levin, Differential neutralization efficiency of hemagglutinin epitopes, antibody interference, and the design of influenza vaccines. Proceedings of the National Academy of Sciences 106, 8701–8706 (2009).

[27] RE Baker, W Yang, GA Vecchi, CJE Metcalf, BT Grenfell, Susceptible supply limits the role of climate in the early SARS-CoV-2 pandemic. Science 369, 315–319 (2020).

[28] NG Davies, et al., Estimated transmissibility and impact of SARS-CoV-2 lineage B.1.1.7 in England. Science 372, eabg3055 (2021).

[29] M Merad, CA Blish, F Sallusto, A Iwasaki, The immunology and immunopathology of covid-19. Science 375, 1122–1127 (2022).

[30] MJ Mina, et al., A Global lmmunological Observatory to meet a time of pandemics. eLife 9, e58989 (2020).

[31] CJE Metcalf, et al., Use of serological surveys to generate key insights into the changing global landscape of infectious disease. The Lancet 388, 728–730 (2016).

[32] CJE Metcalf, MJ Mina, AK Winter, BT Grenfell, Opportunities and challenges of a World Serum Bank - Authors ‘ reply. The Lancet 389, 252 (2017).

[33] J Tang, et al., Respiratory mucosal immunity against sars-cov-2 following mrna vaccination. Science Immunology, DOI: 10.1126/sciimmunol.add4853 (2022).

[34] EJ Topol, A Iwasaki, Operation Nasal Vaccine–Lightning speed to counter COVID-19. Science Immunology 7, eadd9947 (2022).

