## Supplementary Materials for "Medium-term scenarios of COVID-19 as a function of immune uncertainties and chronic disease"


### Framework

#### Model equations

The model equations extend those of [8]. The new state variables are: $I_T$ and $I_Q$ denoting the fraction of individuals with tertiary and quaternary infections, respectively; $S_T$ and $S_Q$ denoting the fraction of individuals with waned susceptibility after a tertiary and quaternary exposure, respectively; $V_S$ and $V_T$ denoting the fraction of individuals vaccinated after a first and second (and beyond) exposure, respectively; and $R_P$, $R_S$, and $R_T$ denoting the fraction of individuals recovered (with complete immunity) from a primary, secondary, and tertiary (and beyond) infection, respectively. Additionally, $\varepsilon_S$, $\varepsilon_T$, and $\varepsilon_Q$ are the relative susceptibility of a secondary, tertiary, and quaternary infection, respectively; $\alpha_S$, $\alpha_T$, and $\alpha_Q$ are the relative transmissibility of a secondary, tertiary, and quaternary infection, respectively.

All other parameters are as in [8]. The (seasonal) transmission rate at time $t$ is denoted $\beta(t)$ (see [27]); the vaccination rate is denoted $\nu$, and $s_{\text{vax}} = 1$ when there is vaccination (otherwise $s_{\text{vax}} = 0$); the recovery rate is denoted $\gamma$; the birth/death rate is denoted $\mu$; and the rates of waning natural and vaccinal immunity are denoted $\delta$ and $\delta_{\text{vax}}$, respectively (and thus the average durations of natural and vaccinal immunity following one exposure are $\frac{1}{\delta}$ and $\frac{1}{\delta_{\text{vax}}}$, respectively).

The model equations are

$$\frac{dS_P}{dt} = \mu - \beta(t)S_P(I_P + \alpha_S I_S + \alpha_T I_T + \alpha_Q I_Q) - \mu S_P - s_{\text{vax}}\nu S_P, \tag{1a}$$

$$\frac{dI_P}{dt} = \beta(t)S_P(I_P + \alpha_S I_S + \alpha_T I_T + \alpha_Q I_Q) - (\gamma + \mu)I_P, \tag{1b}$$

$$\frac{dR_P}{dt} = \gamma I_P - (\delta + \mu + s_{\text{vax}}\nu)R_P, \tag{1c}$$

$$\frac{dR_S}{dt} = \gamma I_S - (\delta + \mu + s_{\text{vax}}\nu)R_S, \tag{1d}$$

$$\frac{dR_T}{dt} = \gamma(I_T + I_Q) - (\delta + \mu)R_T, \tag{1e}$$

$$\frac{dS_S}{dt} = \delta R_P - \varepsilon_S\beta(t)S_S(I_P + \alpha_S I_S + \alpha_T I_T + \alpha_Q I_Q) - \mu S_S + \delta_{\text{vax}}V_P - s_{\text{vax}}\nu S_S, \tag{1f}$$

$$\frac{dS_T}{dt} = \delta R_S - \varepsilon_T\beta(t)S_T(I_P + \alpha_S I_S + \alpha_T I_T + \alpha_Q I_Q) - \mu S_T + \delta_{\text{vax}}V_S - s_{\text{vax}}\nu S_T, \tag{1g}$$

$$\frac{dS_Q}{dt} = \delta R_T - \varepsilon_Q\beta(t)S_Q(I_P + \alpha_S I_S + \alpha_T I_T + \alpha_Q I_Q) - \mu S_Q + \delta_{\text{vax}}V_T - s_{\text{vax}}\nu S_Q, \tag{1h}$$

$$\frac{dI_S}{dt} = \varepsilon_S\beta(t)S_S(I_P + \alpha_S I_S + \alpha_T I_T + \alpha_Q I_Q) - (\gamma + \mu)I_S, \tag{1i}$$

$$\frac{dI_T}{dt} = \varepsilon_T\beta(t)S_T(I_P + \alpha_S I_S + \alpha_T I_T + \alpha_Q I_Q) - (\gamma + \mu)I_T, \tag{1j}$$

$$\frac{dI_Q}{dt} = \varepsilon_Q\beta(t)S_Q(I_P + \alpha_S I_S + \alpha_T I_T + \alpha_Q I_Q) - (\gamma + \mu)I_Q, \tag{1k}$$

$$\frac{dV_P}{dt} = s_{\text{vax}}\nu S_P - \delta_{\text{vax}}V_P - \mu V_P - s_{\text{vax}}\nu V_P, \tag{1l}$$

$$\frac{dV_S}{dt} = s_{\text{vax}}\nu S_S + s_{\text{vax}}\nu R_P + s_{\text{vax}}\nu V_P - \delta_{\text{vax}}V_S - \mu V_S - s_{\text{vax}}\nu V_S, \tag{1m}$$

$$\frac{dV_T}{dt} = s_{\text{vax}}\nu(S_T + S_Q) + s_{\text{vax}}\nu R_S + s_{\text{vax}}\nu V_S - \delta_{\text{vax}}V_T - \mu V_T. \tag{1n}$$

In all our scenarios, we set $\alpha_P = \alpha_S = \alpha_T = \alpha_Q = 1$. We also set $\varepsilon_S = \varepsilon$, $\varepsilon_T = \varepsilon^{2^n}$, and $\varepsilon_Q = \varepsilon^{3^n}$.

#### Calculating Average E and S

We calculate Avg E as follows. First, this quantity is defined for weeks $t$ only when $S_S(t) + S_T(t) + S_Q(t) > 0$, *i.e.* there are some individuals that are susceptible to reinfection. Then,

$$\text{Avg. E}(t) = \frac{\varepsilon S_S(t) + \varepsilon^{2^n} S_T(t) + \varepsilon^{3^n} S_Q(t)}{S_S(t) + S_T(t) + S_Q(t)}.$$

We calculate Avg S as

$$\text{Avg. S}(t) = S_P(t) + \varepsilon S_S(t) + \varepsilon^{2^n} S_T(t) + \varepsilon^{3^n} S_Q(t).$$

#### Modelling clinically important 'Long COVID' cases

To model the fraction of individuals $L$ with a clinically important (Long COVID) case, we assume that a fraction $f_P$, $f_S$, $f_T$, and $f_Q$ of primary, secondary, tertiary, and quaternary infections lead to Long COVID. Furthermore, we assume that the average duration of Long COVID is $\frac{1}{\Phi}$. Thus, we obtain that

$$\begin{aligned}\frac{dL}{dt} &= f_P \beta(t) S_P (I_P + \alpha_S I_S + \alpha_T I_T + \alpha_Q I_Q) + f_S \varepsilon_S \beta(t) S_S (I_P + \alpha_S I_S + \alpha_T I_T + \alpha_Q I_Q) \\ &\quad + f_T \varepsilon_T \beta(t) S_T (I_P + \alpha_S I_S + \alpha_T I_T + \alpha_Q I_Q) + f_Q \varepsilon_Q \beta(t) S_Q (I_P + \alpha_S I_S + \alpha_T I_T + \alpha_Q I_Q) - \Phi L.\end{aligned} \tag{2}$$

We further assume that $f_S = p f_P$, $f_T = p f_S = p^2 f_P$, and $f_Q = p f_T = p^2 f_S = p^3 f_P$. We set $f_P = 0.3$, and vary $p$ for optimistic and pessimistic scenarios, and we set $\frac{1}{\Phi} = 1$ year.

### Accumulation of immunity, total cases, and clinical severity

In Figure S1, we examine the combined effects of immunity accumulation, relative susceptibility to secondary infection, and decreases in severity due to prior exposure on the number and severity of infections. In the more pessimistic - but arguably likely - situation where the duration of complete natural and vaccinal immunity is short (*top row, left panel*, Fig. S1) and the relative

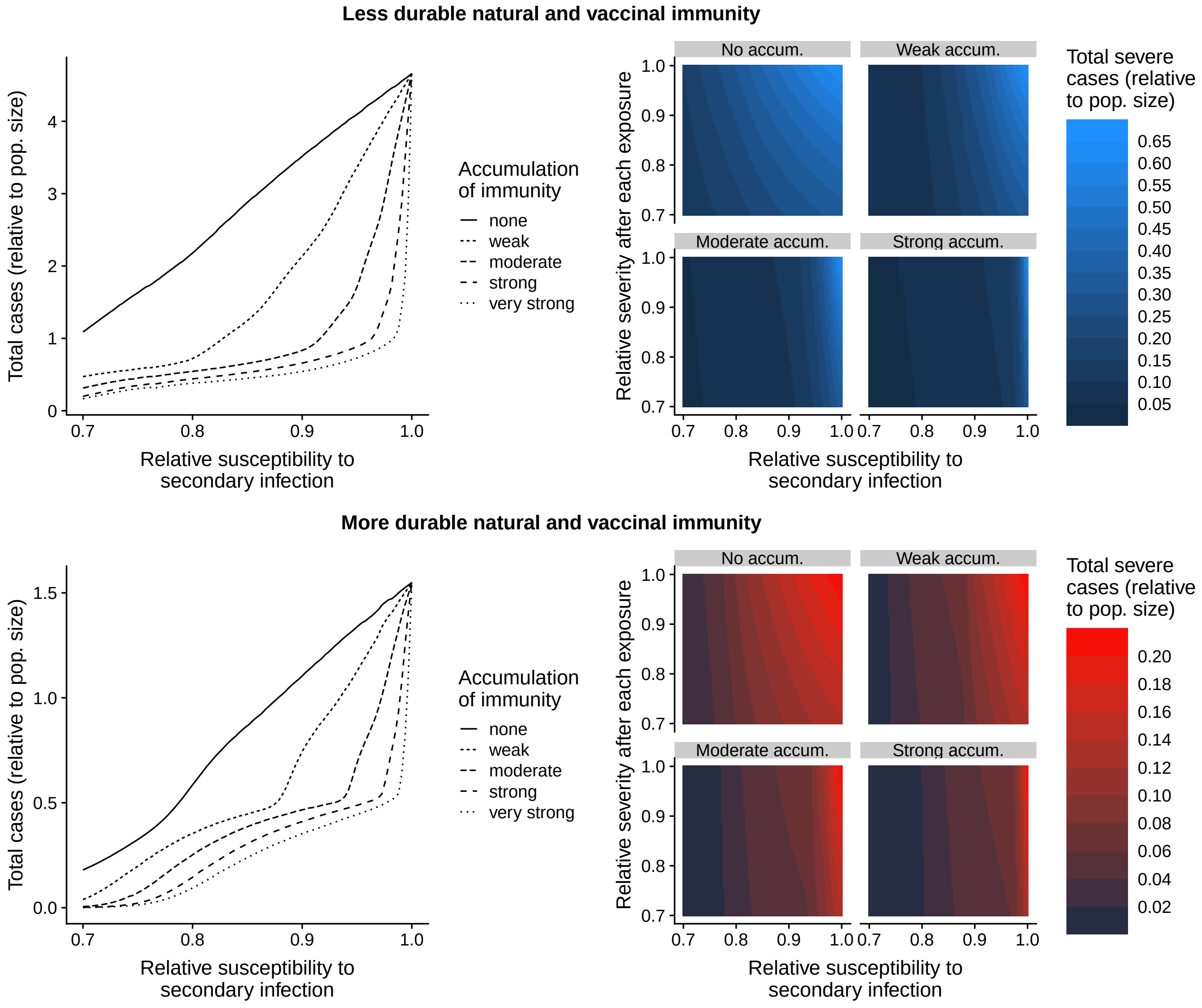


**Figure S1: Total and severe cases (after week 52 and up to week 260) as a function of the relative susceptibility to secondary infection and the accumulation of immunity, for (*Top row*) less durable natural and vaccinal immunity $\left(\delta = \frac{1}{0.25(52)}, \delta_{\text{vax}} = \frac{1}{0.33(52)}\right)$ and (*Bottom row*) more durable natural and vaccinal immunity $\left(\delta = \frac{1}{2(0.25(52))}, \delta_{\text{vax}} = \frac{1}{2(0.33(52))}\right)$. In the right panel of each row, the total severe cases are also a function of the change in severity after each exposure.**

susceptibility to secondary infection is high, individuals are expected to experience multiple infections in the medium-term (*top row, left panel*, Fig. S1). Furthermore, either an increase in accumulation of immunity or a decrease in relative susceptibility to secondary infections tempers the fraction of total cases (*top row, left panel*, Fig. S1), and these two characteristics of host immune responses act synergistically to reduce infections. Finally, as long as some immunity accumulates after subsequent exposures, the rapidity of this accumulation becomes increasingly irrelevant as the relative susceptibility to secondary infection decreases.

Next, in order to examine the potential range of clinical outcomes, we assume that each exposure leads to host immune responses that decrease the subsequent likelihood of experiencing a severe infection (up to quaternary infections). If there is no or weak accumulation of transmission-blocking immunity against infection, then the degree to which a prior exposure protects against severe disease can have a substantial effect on the number of severe infections, particularly when the relative susceptibility to secondary infection is high (*top row, right panel*, Fig. S1). If transmission-blocking immunity accumulates more strongly, then protection against severe disease becomes increasingly less important (*top row, right panel*, Fig. S1). Intuitively, this is due to transmission-blocking immunity limiting infections and thus decreasing the overall potential for severe disease.

Finally, we contrast the above results with epidemiological outcomes in a more optimistic situation with longer-lasting complete natural and vaccinal immunity (*bottom row*, Fig. S1). In this setting, we find that the number of infections (and those that are clinically relevant) are substantially lower than when the duration of immunity is shorter. However, the accumulation of immunity and the relative susceptibility to secondary infection remain important determinants of epidemiological outcomes.
